# The locus coeruleus shows a spatial pattern of structural disintegration in Parkinson’s disease

**DOI:** 10.1101/2021.09.01.21262920

**Authors:** Christopher F. Madelung, David Meder, Søren A. Fuglsang, Marta M. Marques, Vincent O. Boer, Kristoffer H. Madsen, Esben T. Petersen, Anne-Mette Hejl, Annemette Løkkegaard, Hartwig R. Siebner

## Abstract

**Background:** Parkinson’s disease (PD) leads to a loss of neuromelanin positive, noradrenergic neurons in the locus coeruleus (LC) which has been implicated in non-motor dysfunction. “Neuromelanin sensitive” magnetic resonance imaging (MRI) has emerged as a promising tool for mapping the structural integrity of LC in vivo.

**Objectives:** To identify spatial patterns of structural LC disintegration in PD and regions in the LC where structural disintegration is associated with specific non-motor dysfunctions.

**Methods:** 42 patients with PD and 24 age-matched healthy volunteers underwent ultra-high field MRI of the LC using a “neuromelanin sensitive” magnetization transfer weighted (MTw) sequence. The contrast-to-noise ratio of the MTw signal (CNR_MTw_) served as an estimate of structural integrity, slice- and voxel-wise analyses of CNR_MTw_ were performed to map the spatial pattern of structural disintegration, complemented by Principal Component Analysis (PCA). We also tested for correlations between CNR_MTw_ and the severity of non-motor symptoms.

**Results:** Mean CNR_MTw_ of LC was reduced in patients relative to controls. The attenuation of CNR_MTw_ was not uniformly expressed in LC, but confined to the middle and caudal LC. CNR_MTw_ attenuation in caudal LC scaled with the orthostatic drop in systolic blood pressure and apathy ratings. PCA identified a bilaterally expressed component that was more weakly expressed in patients. This component was characterized by a gradual change in CNR_MTw_ along the rostro-caudal and dorso-ventral axes of the nucleus. The individual expression score of this component reflected the overall severity of non-motor symptoms.

**Conclusion:** PD related structural disintegration of LC mainly affects its caudal part and may determine the individual expression of specific non-motor symptoms such as orthostatic dysregulation or apathy.

## (2) Introduction

Parkinson’s disease (PD) is not a mere motor disorder, but leads to a wide range of non-motor symptoms, including sleep disturbances, depression, anxiety, apathy, cognitive impairment and autonomic dysfunction (e.g. orthostatic hypotension, constipation, urinary incontinence) (1). Of these, orthostatic hypotension is a prevalent symptom affecting between 30 and 58% of patients with PD (2). Apathy and depressive symptoms are also frequent in PD with a prevalence of approximately 40% and 35%, respectively (3,4). The pathophysiological mechanisms underlying apathy and depression in PD likely involve multiple neuromodulatory systems including the locus coeruleus (LC) (5,6). Noradrenergic neuromodulation from LC neurons plays a role in the regulation of sleep and arousal, executive function, memory consolidation, motivation and decision making as well as autonomic function (7,8). Accordingly, neurodegeneration of LC noradrenergic neurons in PD has been linked to non-motor symptoms such as REM-sleep Behavior Disorder (RBD), mild cognitive impairment, depression and orthostatic hypotension (9–14).

The importance of LC neurodegeneration in the symptomatology of PD is emphasized by the idea that LC neurons show progressive accumulation of Lewy bodies composed of misfolded alpha-synuclein (15,16). The LC is a paired, elongated nucleus in the dorsolateral pontine tegmentum consisting of approximately 50.000 pigmented noradrenergic neurons (17,18). The nucleus extends from the inferior colliculus rostrally to the lateral face of the 4th ventricle caudally occupying approximately 13 mm^3^ (18). For many decades, the LC was viewed as a unitary nucleus that processes the same type of information across its structure, distributing a uniform message across its efferent projections (7,19). However, recent research has shown a topographical organization of neurons based on target structures with forebrain projecting neurons located rostrally and neurons projecting to the basal ganglia, cerebellum and spinal cord located in the middle and caudal parts of the nucleus (20,21) with corresponding different functional modules across the structure (7). Yet, there is to date little information on whether neurodegeneration in PD affects the LC in a uniform manner.

In recent years, neuromelanin sensitive magnetic resonance imaging (MRI) has been introduced as a sensitive means of assessing PD related changes in the substantia nigra and locus coeruleus (22–25). On neuromelanin MRI the LC is visible as a paired, elongated, hyperintense structure along the floor of the fourth ventricle in the upper pons. The voxel intensity has been shown to be closely associated with the number of neuromelanin-rich neurons in the LC (26). In the midbrain, the substantia nigra can be visualized as a hyperintense structure on neuromelanin MRI, and the voxel intensity has been found to correlate with neuromelanin concentration in the substantia nigra (27). Recently, magnetization transfer weighted (MTw) imaging has been introduced providing increased LC contrast-to-noise ratio (CNR) compared to conventional turbo spin echo sequences (28). A recent study investigating LC integrity in PD using MTw MRI showed spatial inhomogeneity in the pattern of neurodegeneration with a decrease in CNR in the caudal part of the nucleus, but not in the middle or rostral parts (25).

Here, we used ultra-high field (7 tesla) MTw MRI to investigate the extent and pattern of LC neurodegeneration in PD at high spatial resolution and its relationship with non-motor symptoms. We hypothesized that patients would show a reduced mean MTw signal in the LC reflecting the neuronal loss of noradrenergic neurons. We expected the signal reduction in the LC would scale with the severity of non-motor symptoms. Furthermore, we explored the spatial resolution of 7 tesla MRI to identify spatial gradients of structural disintegration and how they are related to non-motor symptoms.

## (3) Methods

### Participants

We recruited 49 patients with PD and 27 healthy, age-matched controls (HC) as part of a larger study investigating brainstem changes in PD. Seven PD patients were excluded due to contraindications to MRI (3), unreported neurological conditions (1), incidental findings (1) and poor scan quality (2). Three HC were excluded due to unreported neurological conditions (2) and poor scan quality (1). Patients with PD were recruited from the Movement Disorders outpatient clinic at the Department of Neurology, Copenhagen University Hospital Bispebjerg (Copenhagen, Denmark) and private practice neurology clinics based in the Copenhagen Region. All participants in the PD group were required to have a clinical diagnosis of PD assessed by a neurologist and were additionally required to meet the MDS Clinical Diagnostic Criteria for “Clinically Established PD” or “Clinically Probable PD” (29). Exclusion criteria were pregnancy or breastfeeding, history of other neurologic or psychiatric disease, pacemaker or other implanted electronic devices and claustrophobia. The PD group spanned a large range in terms of disease duration (0-17 years) and time since symptom onset (1-18 years) and included patients with mild to moderate disease (Hoehn & Yahr stages 1 - 3).

Healthy control participants were recruited by online advertisements on www.forsoegsperson.dk and were required to be 18 years or older with no history of neurologic or psychiatric disease and fulfill none of the exclusion criteria mentioned above.

The study was approved by the Regional Committee on Health Research Ethics of the Capital Region of Denmark (Record-id: H-18021857). All participants gave their written informed consent to participate in the study. The study was pre-registered at ClinicalTrials.gov [Identifier: NCT03866044].

### Study procedures

Participants underwent a neurological examination to exclude participants that exhibited symptoms of any neurological condition other than PD. Patients’ motor and non-motor symptom severity was assessed in the ON-state (on their usual dopaminergic treatment) using the Unified Parkinson’s Disease Rating Scale (UPDRS) and the Non-Motor Symptom Scale (NMSS) (30,31). Patients’ blood pressure was measured in the supine position following a 5 minute rest period after which a series of consecutive measurements were made over a period of 3 minutes upon standing up (32). The maximum change in systolic blood pressure was recorded as a measure of orthostatic hypotension. Apathy and depression were assessed in all participants using the Lille Apathy Rating Scale (LARS) and Beck’s Depression Inventory-II (BDI-II) (33–36). Additionally, we registered age and sex, as well as patients’ disease duration, time since onset of motor symptoms and medication status.

### Magnetic resonance imaging data

Structural MRI data was collected with a Philips Achieva 7T scanner (Philips, Best, The Netherlands) equipped with a 32-channel Nova head coil (Nova Medical,Inc., MA, USA). We scanned patients in the ON-medication state to limit tremor-related movement while inside the scanner. As an anatomical reference to guide coregistration and normalization, we acquired T1w, high-resolution (1 mm isotropic) Magnetization Prepared RApid Gradient Echo (MPRAGE) images with a field-of-view covering the entire head (200×288×288 voxels), echo time / repetition time = 2.2 / 4.9 ms, acquisition time = 3:30 minutes. To assess the integrity of the LC, we acquired MT-weighted images using a 3D high-resolution (voxel size 0.4×0.4×1.0 mm) ultra fast gradient echo sequence aligned to the AC-PC line with a field-of-view covering the midbrain and rostral pons (640×640×34 voxels), echo time / repetition time = 4.1 / 8.1 ms, flip angle = 7 degrees, 2 averages. MT-saturation was achieved by applying 16 block-shaped pre-pulses at a frequency offset of 2 kHz (flip angle = 278 degrees, duration = 10 ms), acquisition time = 8 minutes. For coregistration purposes, we also acquired T1-weighted images with identical acquisition parameters to the MT-weighted sequence, but with a 0 degree flip angle for the off-resonance pre-pulses.

### Image post processing

Post processing of imaging data and calculation of CNR maps was performed following a procedure comparable to that described in a recent study of the LC in healthy participants (37).

T1-weighted anatomical reference images were upsampled to a resolution of 0.5 mm isotropic. Upsampled T1-weighted reference images and high resolution MT-weighted and T1-weighted images were corrected for low-frequency spatial intensity inhomogeneities using the Advanced Normalization Tools (ANTs) software (N4 bias field correction, 5 resolution levels, number of iterations per level: 50×50×30×20, convergence threshold: 1×10-6, isotropic sizing for b-spline fitting: 200) (38).

Within-subject co-registration and subsequent normalization to template space was performed using ANTs by rigid-body, affine and non-linear registration using the following procedure. Within-subject co-registration was achieved by rigid-body registration of the high resolution T1-weighted images to the T1-weighted reference scan and by rigid-body registration of the high resolution MT-weighted images to high resolution T1-weighted images and subsequently to the T1-weighted reference scan. High resolution images were transformed to individual subjects’ T1-weighted space for evaluation of registration accuracy.

T1-weighted reference images were normalized to the MNI 0.5 mm ICBM152 (International Consortium for Brain Mapping) T1w asymmetric template for analysis (39) by rigid-body, affine and non-linear registration. Finally, MT-weighted and T1-weighted images were transformed to MNI template space by combining the transformation parameters obtained from within-subject co-registration with the transformation parameters and warp-field obtained from the normalization step. Contrast-to-noise-ratio (CNR_MTw_) maps were calculated from normalized MT-weighted images by subtracting the mean signal intensity of a 4 × 4 × 4 mm cubic reference region placed centrally in the pontine tegmentum (PTref) and dividing by the standard deviation of this reference region: CNR_MTw_ = (SI - mean_PTref_) / sd_PTref_. We chose to quantify LC contrast relative to the standard deviation of the reference region instead of the mean signal, as the standard deviation provides a better estimate of the noise in the imaging data (37).

A region of interest (ROI) for the LC was defined based on a probabilistic LC atlas constructed from 7T MT-weighted images from 53 healthy participants (age range 52-84 years, mean: 66 years) (37). The liberal version of this atlas (5% probability) was chosen as ROI to allow for interindividual difference of the spatial extent of the LC. Visual inspection of the atlas overlaid on an average MTw image of all 24 healthy participants in this study revealed that for our data the atlas also included hyperintense voxels forming the border of the 4th ventricle. This hyperintense border would cause erroneously high CNR-values if included in the region of interest, because it does not correspond to the anatomical location of the LC. Therefore, we created a binary mask applying threshold-based segmentation of the brainstem using the MNI 0.5 mm ICBM152 (International Consortium for Brain Mapping) T1w, non-linear, asymmetric template. All voxels outside the segmented brainstem were masked out, which effectively excluded the hyperintense border voxels (Supplementary figure 1).

As an overall index of LC integrity, mean CNR_MTw_ was extracted from the bilateral LC ROI as well as from every slice from the rostral to the caudal most extent of the LC ROI in the z-direction for each side separately.

### Statistical analyses

Statistical analyses were performed using R (R Core Team 2019, v4.0.4) and Python (Python Software Foundation, v. 3.7.9). Between group differences were tested using independent samples t-tests of mean CNR_MTw_ values of right and left LC, mean CNR_MTw_ values of left & right LC per slice and single-voxel CNR_MTw_ values.

We used one-tailed correlation analyses to test our hypotheses, that LC CNR would correlate negatively with overall non-motor symptom severity (NMSS), apathy (LARS) and depression (BDI-II), and positively with orthostatic drop in systolic blood pressure in patients with PD. Correlations were assessed for the mean CNR_MTw_ of both LC, slicewise and voxelwise CNR_MTw_-values to explore at which rostro-caudal levels CNR_MTw_ correlated with non-motor symptom severity. One-tailed Pearson’s product moment correlation coefficient and one-tailed Spearman’s rho was calculated as appropriate. Statistical significance was accepted at a threshold of *p* < 0.05 family-wise error corrected.

Cluster-level inference was used to explore the spatial extent of between group differences and correlations with non-motor symptom severity. To this end, we applied threshold-free cluster enhancement (E=0.5, H=2 and 6 neighborhood connectivity) with 10.000 permutations to correct for multiple comparisons and included age as a regressor of no interest (40).

We further hypothesized that LC degeneration may manifest itself in distributed patterns (e.g. spatial gradients) across LC. In an exploratory analysis, we identified distributed patterns in the CNR_MTw_-maps over the LC search space using Principal Component Analysis (PCA). To this end, we extracted CNR_MTw_-maps from all subjects, z-scored the maps across subjects on a voxel-by-voxel basis and applied PCA across subjects. We focused on the three first components that explained most variance in the data (52.7 % variance explained). We assessed between-group differences in principal component (PC) loadings for each PC using two-tailed independent samples t-tests and investigated if the loadings significantly correlated with clinical scores in the PD group using two-tailed Spearman Rank correlations.

## (4) Results

### Clinical data

The PD group and control group did not differ in age (Z = 548, p = 0.557) or gender (χ^2^ = 0.471, p = 0.493) (Table 1). Patients had significantly higher BDI-II scores (Z = 263.5, p = 0.001) and LARS scores (t(56.6) = -2.26, p = 0.028) compared to healthy volunteers. 17 out of 42 patients showed orthostatic hypotension, i.e. a systolic pressure drop of ≥20 mmHg or diastolic pressure drop of ≥10 mmHg, as defined by the Consensus Committee of the American Autonomic Society and the American Academy of Neurology. Two healthy participants were treated with antihypertensive treatment in the form of beta-1-receptor antagonists and nine patients were on some form of antihypertensive treatment including beta-1-receptor antagonists (N = 4), diuretic (N = 3) and ACE-inhibitors or calcium channel blockers (N = 3) as either monotherapy (N = 2) or in combination (N = 7).

**Table 1.** Demographics of the study population.

|  | Healthy<br>(N=24) | Parkinson<br>(N=42) | *p* |
| --- | --- | --- | --- |
| Sex (male/female) | 16/8 | 23/19 | 0.493 |
| Age (years) | 71.5 (43.0, 80.0) | 67.5 (38.0, 84.0) | 0.557 |
| Disease duration<br>(years) | - | 3.66 (0.250, 17.3) | - |
| Time since symptom<br>onset (years) | - | 6.06 (0.980, 18.3) | - |
| Modified Hoehn &<br>Yahr stage | - | 2 (1, 3) | - |
| Levodopa equivalent<br>dose (mg/day) | - | 479 (100, 2140) | - |
| Most affected side<br>(right/left) | - | 17/25 | - |
| UPDRS-III | - | 23.4 ± 9.27 | - |
| NMSS | - | 31 (3, 118) | - |
| MoCA | 28 (23, 31) | 28 (17, 30) | 0.371 |
| BDI-II | 3 (0, 11) | 8 (0, 20) | 0.001 |
| LARS | -25.5 ± 4.93 | -22.4 ± 6.09 | 0.028 |

*Data given as mean ± SD or median (range). Between group differences was assessed using student's t-test for normally distributed data and Wilcoxon rank-sum test when normal distribution could not be assumed. Between group difference in gender distribution was assessed using Pearson's Chi-squared test with Yates' continuity correction. UPDRS-III = Unified Parkinson's Disease Rating Scale, subscore III, NMSS = Non-Motor Symptom Scale Score, MoCA = Montreal Cognitive Assessment, BDI-II = Beck's Depression Inventory.*

### Overall MTw contrast in the locus coeruleus

Patients had lower mean CNR_MTw_ values in the right LC than age-matched healthy volunteers (t(57.2)= -2.96, p = 0.002), confirming disease-related structural disintegration of the LC (Figure 1A). However, left LC CNR_MTw_ values were not significantly lower in patients compared to healthy volunteers (t(62.7) = -0.69, p = 0.25). We found no significant correlations between mean CNR_MTw_ in LC and the severity of overall non-motor symptoms (NMSS) or depression (BDI-II). For the right LC, there was a trend towards patients with low mean CNR_MTw_ having higher apathy ratings, however this correlation was not significant following multiple comparisons correction. Mean CNR_MTw_ values in the left LC correlated positively with orthostatic change in systolic blood pressure. Patients with low mean CNR_MTw_ values in the left LC exhibited larger systolic blood pressure decreases than patients with high mean CNR_MTw_ values (rho = 0.42, uncorrected p = 0.003, FWE corrected p = 0.037) (Figure 1C). A similar trend was observed for the right LC, however the association was not significant following multiple comparisons correction. The inverse relation between mean CNR_MTw_ values in the left LC and orthostatic drop in systolic blood pressure was still significant after exclusion of patients receiving antihypertensive treatment (Supplementary figure 3D).

**Figure 1.**
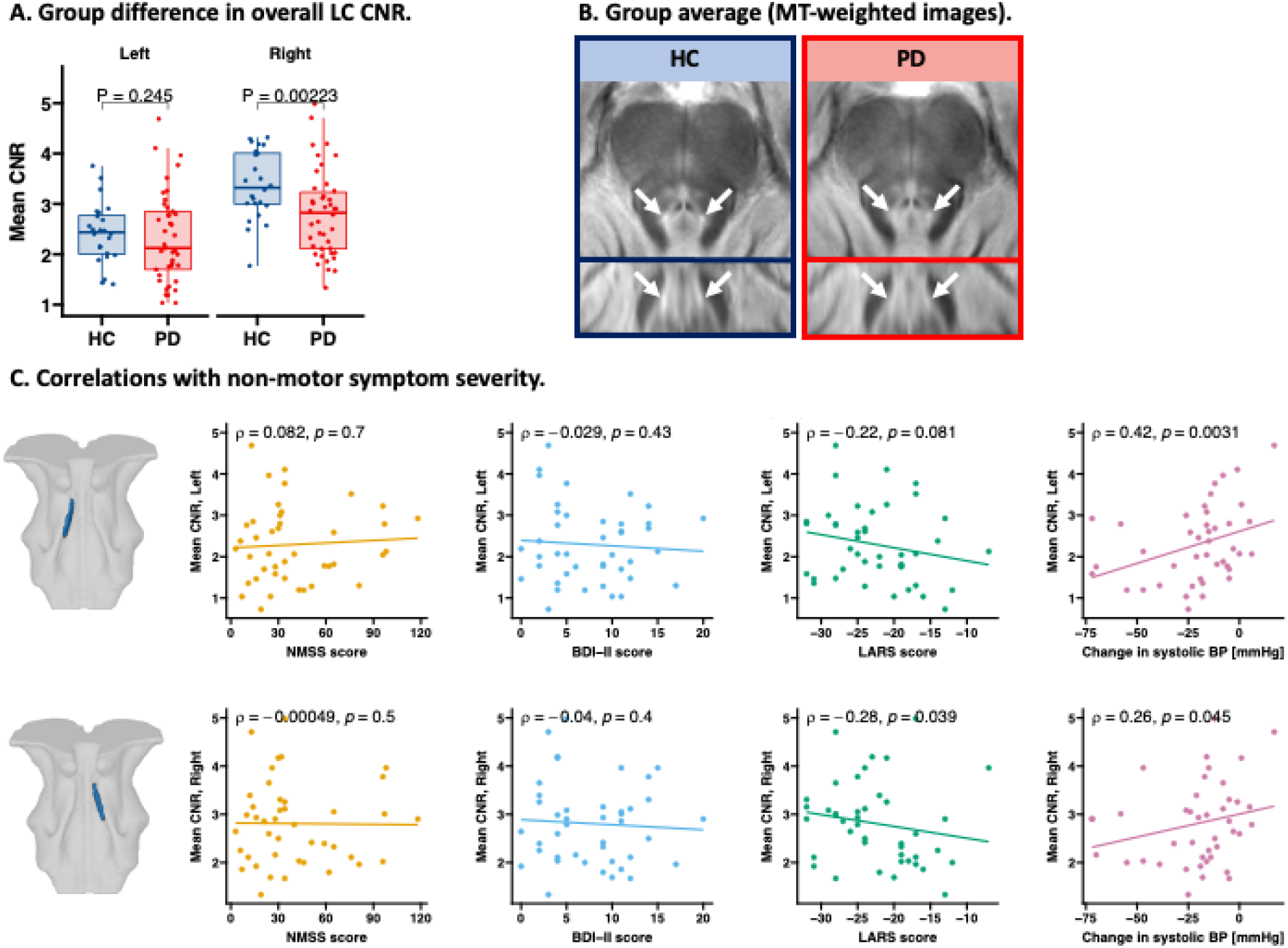
Overall group difference in LC CNR_MTw_ & correlations with non-motor symptom severity. Patients had lower mean CNR_MTw_ in the right LC compared with age-matched healthy volunteers (t(57.2)= -2.96, p = 0.002) (A). The reduction in CNR_MTw_ is visualized in group average MTw maps (B). Mean CNR_MTw_ of the left LC orrelated with othostatic change in systolic blood pressure. No significant correlations were found for overall non-motor symptom severity (Non-motor Symptom Scale; NMSS), depressive symptoms (Beck’s Depression Inventory II; BDI-II score) or apathy (Lille Apathy Rating Scale; LARS) following multiple comparisons correction (C).

MRI studies have shown considerable age-related variability of neuromelanin signal in healthy patients (7,41,42). We therefore performed an explorative correlational analysis which included patients and healthy controls for those variables, that had been collected in both groups. Combining the two groups revealed a significant negative correlation between LC CNR_MTw_ and apathy (rho = -0.29, p = 0.0084), but not depressive symptoms (rho = -0.17, p = 0.081) (Supplementary figure 2). NMSS scores and orthostatic change in systolic blood pressure had not assessed in healthy controls. No significant correlations were found between overall LC CNR_MTw_ values and other clinical variables such as disease duration, time since symptom onset and levodopa equivalent dose (Supplementary figure 3A & B).

### Changes in MTw contrast of the locus coeruleus along its rostro-caudal axis

We hypothesized that structural disintegration is not homogenously expressed in the LC. This hypothesis was tested in two complementary analyses, testing for between-group differences at the slice and voxel level. Slice-wise comparison revealed that PD patients had lower mean CNR_MTw_ in the caudal part of LC than healthy controls (Figure 2A). The between-group difference was significant for slice-wise CNR_MTw_ values in the right middle and caudal LC, including slices at z-coordinates 91 to 102, FWE-adjusted p < 0.05 (Figure 2B). CNR_MTw_ values from slices in the left caudal LC correlated positively with orthostatic systolic blood pressure change (from z = 86 - 96). A negative correlation between CNR_MTw_ values in the caudal part of the nucleus was also observed for apathy severity, however this finding did not remain significant following multiple comparisons correction (Figure 2C).

**Figure 2.**
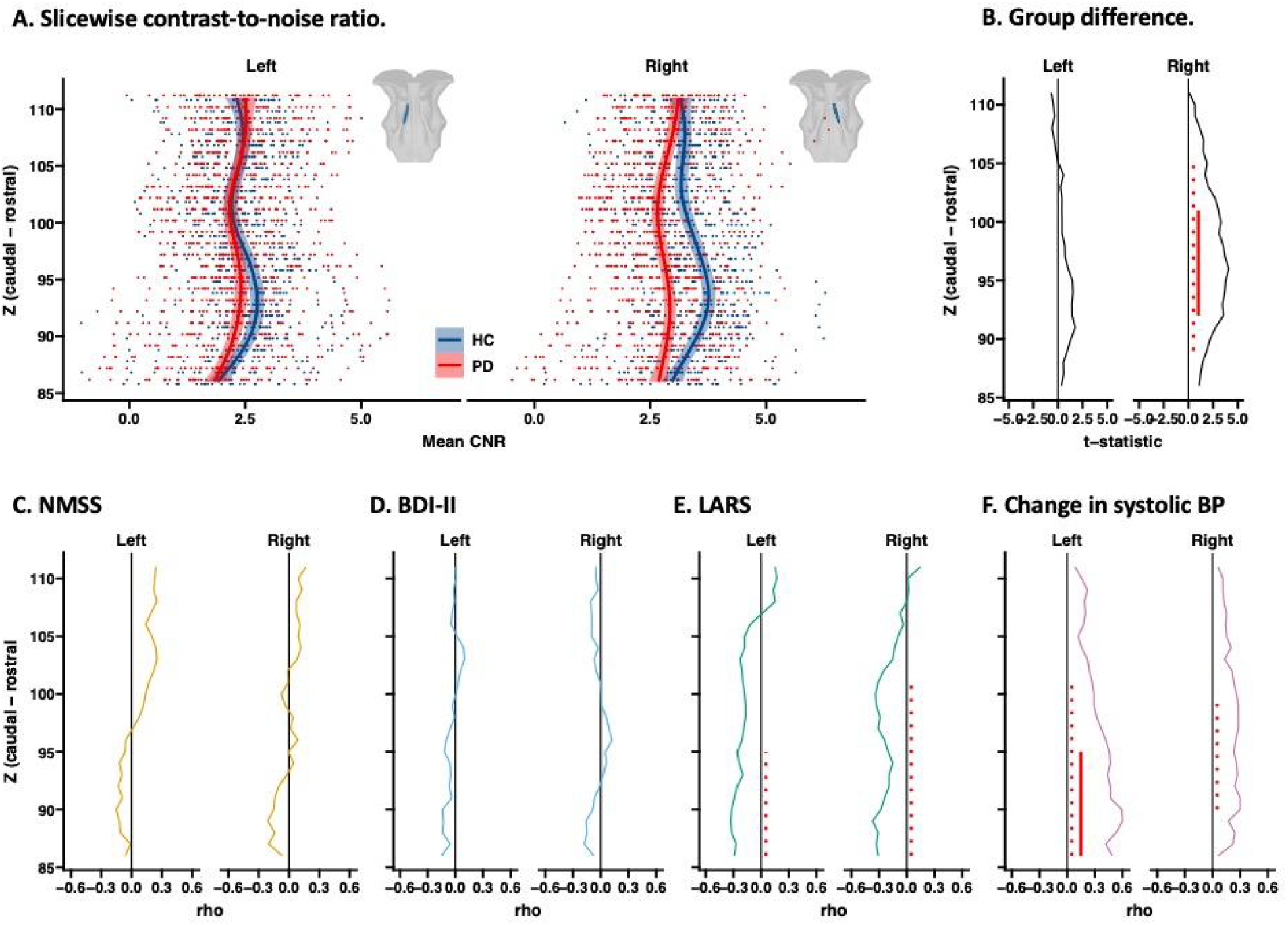
Rostro-caudal, slicewise differences in LC CNR_MTw_ & correlations with non-motor symptom severity. By extracting the slicewise LC CNR_MTw_ along the rostro-caudal extent of the nucleus, we found lower CNR_MTw_ values in the caudal part of the right LC in patients with PD. Groupwise average CNR_MTw_ values are vizualized by fitting a local polynomial regression function (A). Per slice t-statistic and significance level are presented in panel (B) (Red bar indicates slices at which CNR_MTw_ was significantly lower (FWE-adjusted p < 0.05). LC CNR_MTw_ values were found to correlate with apathy (E) and orthostatic hypotension (F). Red bars indicate slices at which CNR_MTw_ correlated with the severity of non-motor symptoms (dashed = unadj. p < 0.05, solid = FWE-adjusted p < 0.05.

Voxel-wise analyses mirrored the findings obtained by slice-wise analysis. Patients showed a cluster of voxels in the right caudal LC with significantly reduced CNR_MTw_ values relative to healthy controls (TFCE, FWE-adjusted p<0.05, cluster size: 98 voxels, Figure 3A). A cluster of voxels in the left caudal LC showed a relationship with orthostatic change in systolic blood pressure (FWE-adjusted p<0.05, cluster size: 106 voxels, Figure 3B). The lower the CNR_MTw_ in these voxels, the higher was the drop in blood pressure when rising up. In a small cluster in the right caudal LC, CNR_MTw_ values were associated with apathy severity as indexed by the LARS-score (FWE-adjusted p<0.05, cluster size: 10 voxels, Figure 3C). No significant clusters were found for general non-motor symptom severity (NMSS) or depressive symptoms (BDI-II).

**Figure 3.**
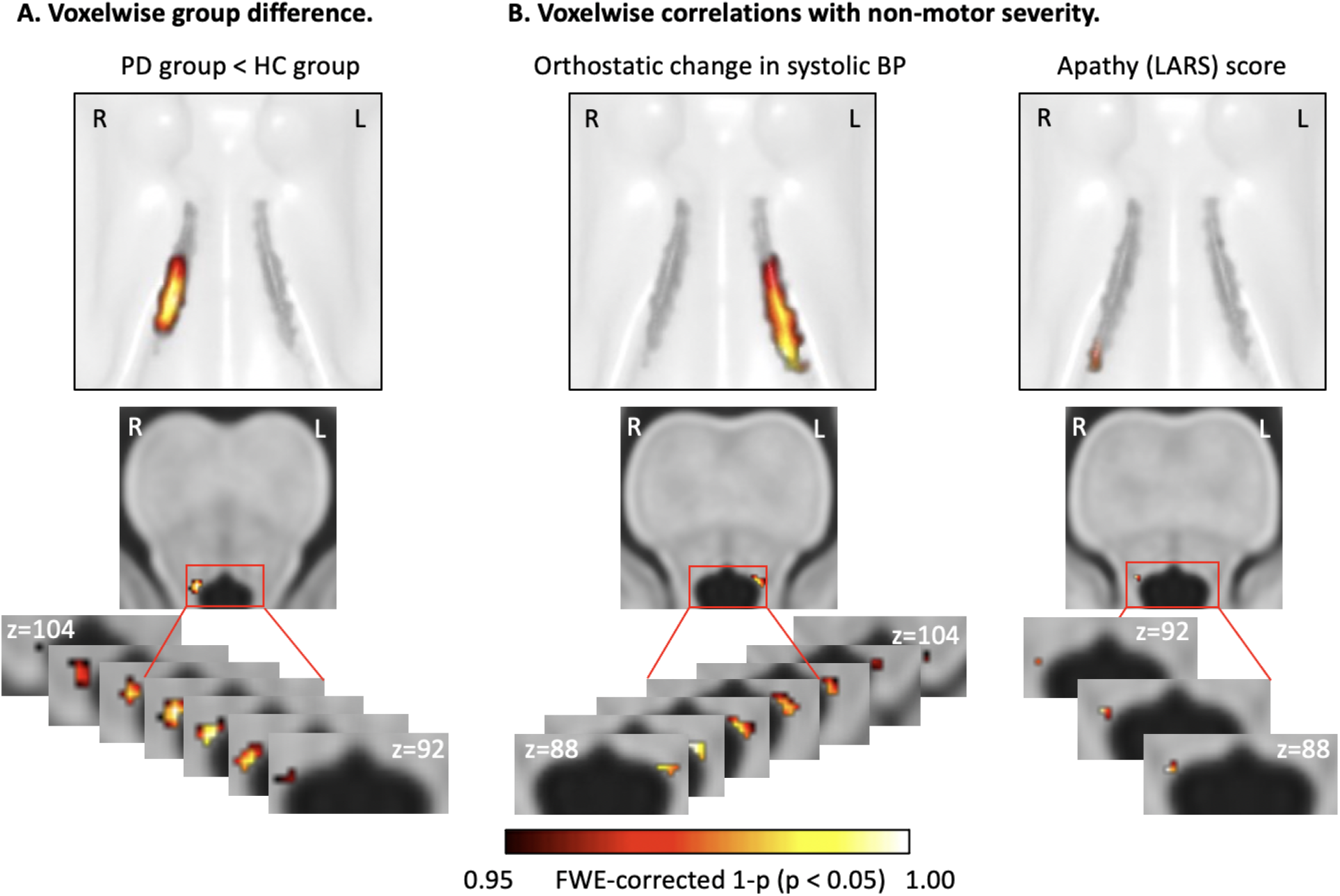
Voxelwise differences in LC CNR_MTw_ & correlations with non-motor symptom severity. Between group difference in LC CNR_MTw_ was assessed voxelwise by applying threshold free cluster enhancement. Patients had reduced CNR_MTw_ in a cluster of voxels in the right caudal LC (A). Voxelwise correlation with non-motor symptom severity was assessed in the PD group. Orthostatic change in systolic blood pressure correlated with CNR_MTw_ values in a cluster in the left caudal LC and apathy correlated negatively in cluster in the right caudal LC (B). Correlations were tested with age as a regressor of no interest. Maps are thresholded at FWE-adjusted p < 0.05. Top panels visualize the locations of significant clusters on interpolated 3D-renderings.

### Principal component analysis

Voxel-wise PCA yielded a set of orthogonal principal components (PCs), with component weights shown in Figure 4B. The weights of PC1 were all positive suggesting a weighted sum of CNR over the entire LC. The spatial pattern of the PC2 component weights corresponded to a combination of rostro-caudal and dorso-ventral gradients within the bilateral nuclei with negative weights in the caudal and dorsal part of the nucleus and positive weights in the rostral and ventral part. We found a significant group difference in PC2 scores with more negative mean scores in the HC group compared with the PD group [F(1,64)=9.192, FWE-adjusted p = 0.0033], consistent with patients having a loss of CNR in caudal parts of the nucleus (Figure 4A). The group difference in PC2 scores was presumably driven by high CNR values in caudal-dorsal voxels relative to rostral-ventral voxels in HC, resulting in negative loadings in healthy controls. In contrast, PD patients exhibited more variable loadings and a shift towards more positive loading values, presumably reflecting an attenuated rostro-caudal gradient. In the PD group, the more positive the loadings of PC2, the higher the were the individual NMSS-scores in patients (rho = 0.33, p = 0.035). A similar trend was observed towards a correlation with LARS score (rho = 0.30, p = 0.053) (Figure 4C).

**Figure 4.**
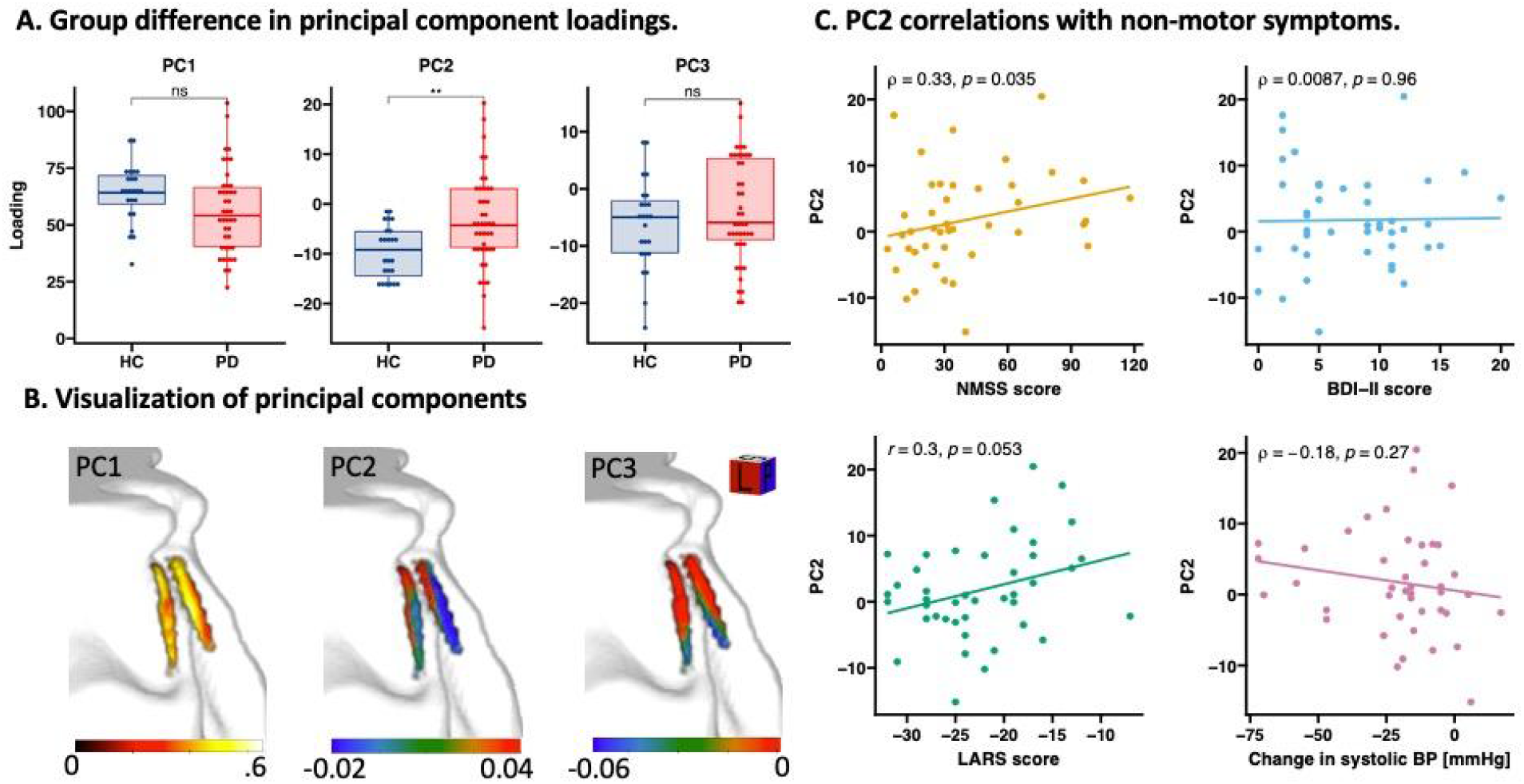
Principal component analysis. Principal Component Analysis was applied on CNR_MTw_ maps from all participants to investigate patterns of disease related changes in LC CNR_MTw_. Loadings on Principal Component 2 reflecting a gradient along the rostro-caudal and dorso-ventral axes of the nucleus were different across groups (A). This gradient (visualized in B) was found to correlate with overall non-motor symptom severity (Non-motor Symptom Scale; NMSS) and a trend was observed towards a correlation with apathy (Lille Apathy Rating Scale; LARS), but not with depression (Beck’s Depression Inventory II; BDI-II score) or orthostatic change in systolic blood pressure (C).

## (5) Discussion

### Summary of main findings

Using ultra-high field MTw MRI, we were able to pinpoint how PD affects the structural integrity of the LC. Our study revealed three main findings. First, mean CNR_MTw_ was reduced in the right LC, confirming a structural disintegration of LC in PD. Second, CNR_MTw_ attenuation was not homogenously distributed in LC, but restricted to its caudal part. Third, the stronger CNR_MTw_ was reduced in the caudal LC, the larger was the orthostatic drop in blood pressure and higher were the individual apathy ratings.

### Overall reduction of the neuromelanin sensitive signal in locus coeruleus

The reduction of mean LC CNR_MTw_ in PD is in agreement with previous MRI studies performed at high field strength, and has also been demonstrated in patients with REM sleep behavioural disorder (RBD) (9,13,14,22,24,43). This robust and replicable reduction of LC contrast on neuromelanin sensitive MR images provides an in-vivo indication of the loss of pigmented LC neurons demonstrated in post-mortem histological studies (16,44–46). Importantly though, in our study as well as previous neuroimaging studies, the attenuation of CNR_MTw_ did not correlate with disease duration or clinical measures of motor severity. As such, the overall reduction in mean CNR_MTw_ does not seem to parallel disease duration or motor related features of PD attributable to dopaminergic neuronal loss and nigrostriatal dysfunction. Similarly, only one post-mortem study has been able to demonstrate an association between LC neuronal loss and disease duration (47), while 16 studies did not report or find any associations with disease duration or severity of motor symptoms (reviewed in (46)). Therefore, we conclude that neuromelanin sensitive MRI of the LC captures an aspect of PD related neurodegeneration, which does not scale with disease duration or motor severity. As outlined in more detail below, we argue that neuromelanin sensitive MRI of LC probes neurodegenerative processes in LC that confer non-motor symptoms.

### Spatial gradient of signal loss in the locus coeruleus

Both voxelwise and slice-wise analyses showed a clear rostro-to-caudal gradient in CNR_MTw_. The decrease in CNR_MTw_ was confined to the caudal portion of the LC, especially on the right side relative to healthy age-matched controls. This is in agreement with a recent high-field MRI study in which LC contrast was attenuated in PD patients in the middle and caudal parts of the LC, but not the rostral part (48). A recent ultra-high field MRI study reported the same spatial attenuation of CNR_MTw_ in slices and voxels of the caudal part of the LC (49). Complementing our slice-wise and voxel-wise analyses, PCA identified a bilaterally expressed spatial component of CNR_MTw_ values that was more weakly expressed in patients.

This component was characterized by a gradual change in CNR_MTw_ along the rostro-caudal and dorso-ventral axes of the nucleus. Together, these findings provide converging evidence that the PD related structural alterations revealed by neuromelanin sensitive MRI are more pronounced in caudal parts of the LC.

Our finding that structural disintegration of the LC is mostly present in the caudal part is in apparent contrast to post-mortem studies, which reported a uniform neuronal loss over the entire LC with PD and a preferential degeneration of rostral LC in Alzheimer’s disease (44,45,50–52). A post-mortem study in seven patients with PD and three healthy control cases found similar relative cell loss in rostral, middle and caudal LC sections (45), yet the greatest absolute cell loss was noted in the middle section. Another post-mortem study in six patients with PD and seven healthy control cases reported that the magnitude of LC cell loss analyzed as the percent loss from normal did not differ in rostral and caudal portions (44). However, a closer evaluation of that data indicates a more pronounced cell loss in caudal compared with rostral sections. In support of this, a post-mortem study in 11 patients with PD found degenerative changes in the entire LC, but most severely in caudal and middle segments (47). More post-mortem studies are needed to address this apparent discrepancy. It might be possible that CNR_MTw_ is not only affected by the magnitude of cell loss but also by dysfunctional neuromelanin positive cells that have not yet undergone apoptosis.

### Structure-function relationship

The clusters in caudal LC showing the strongest attenuation in CNR_MTw_ coincided with the clusters that showed a structure-dysfunction relationship for specific non-motor symptoms. We found no correlations with overall non-motor symptom severity, cognitive performance or depressive symptoms. However, the regional attenuation of the neuromelanin signal in the caudal portion of the right LC scaled with the severity of apathy, while regional attenuation of the neuromelanin signal in the left caudal LC predicted the magnitude of systolic blood pressure drop during orthostatic challenge. The latter correlation was evidenced by slice-wise and voxel-wise analyses.

The demonstration of an association between neuromelanin MRI contrast of LC and specific non-motor symptoms in PD extends previous MRI work showing a link between the neuromelanin sensitive MRI signal in LC and RBD, mild cognitive impairment and depressive symptoms (10,12,12,13). Our finding that a low CNR_MTw_ in right caudal LC scales with the individual expression of apathy confirms and extends a recent MTw 7T MRI study, showing a correlation between LC integrity and apathy in a pooled cohort of patients with PD and progressive supranuclear palsy (49). Together, these findings support a role of noradrenergic dysfunction in the development of apathy in PD which needs to be elucidated in future studies.

The positive correlation between attenuated CNR_MTw_ in the left caudal LC and orthostatic drop in systolic blood pressure in patients with PD is a novel finding, extending previous lines of research into noradrenergic dysfunction in PD. This notion is supported by a recent PET study, which found that the distribution volume ratio of the noradrenaline transporter PET-tracer 11C-MeNER in the LC and hypothalamus correlated with orthostatic change in systolic blood pressure, but only when including healthy participants in the analysis (13). In support of our finding, a neuromelanin sensitive MRI study performed in healthy participants found a negative correlation between LC contrast and high frequency heart rate variability especially in older adults (53). The association of loss of LC integrity, noradrenergic dysfunction and orthostatic hypotension and abnormal heart rate variability could be explained by a loss of LC mediated sympathetic inhibition of the parasympathetic modulation of the heart by the vagus nerve (54–56). Co-occurrence of degeneration of LC neurons and peripheral postganglionic sympathetic neurons (57,58) could also explain the association with orthostatic hypotension. Taken together, these findings support the notion that noradrenergic neurodegeneration in LC makes a critical contribution to the development of orthostatic hypotension in PD.

### Strengths and limitations

This study has several strengths. To improve the accuracy of our CNR measurements, we acquired MT-weighted images with a high in-plane spatial resolution to limit partial volume effects, applied a T1-driven normalization approach, which was independent of the neuromelanin contrast we wished to quantify and extracted CNR values using a pre-existing LC atlas constructed from a similar dataset and a similar central pontine reference region. Minimizing any form of manual interaction with the data, we applied a computationally intensive co-registration and normalization procedure that has previously been reported to allow for accurate co-registration and normalization, and visually verified that normalization results were accurate (37). This is highly important as LC is a small nucleus and even minor normalization and co-registration errors can introduce undesired variability in the results. We also excluded hyperintense voxels at the border of the fourth ventricle to obtain accurate contrast measurements and ultimately to enable the assessment of PD-related changes in different parts of the LC.

The study also has some limitations. The neurobiological mechanisms contributing to the neuromelanin MRI contrast is a matter of ongoing debate. Magnetization transfer effects have been proposed to be the main source of contrast, but a contribution of T1 shortening from neuromelanin-iron compounds has also been suggested (28,59,60). Further, MTw MRI is not specific to neuromelanin rich structures, and hyperintensity is also observed in the periaqueductal grey, which has low neuromelanin content (27). Our MT-weighted acquisition had limited spatial resolution in the slice direction, which could lead to partial volume effects in extreme ends of the nucleus. CNR was consistently higher in the right LC compared to the left and mean *CNR*_*MTw*_ values were only significantly lower in the right LC in patients. The notion of a right-left asymmetry is not supported by post-mortem studies (45,61–63). Spatial variations in the B1-field between vendors may account for this apparent asymmetry. Previous neuroimaging studies have found a similar asymmetry with higher right LC contrast in studies using Philips scanners and higher left LC contrast in studies using Siemens scanners (42,60,64,65). This right-left asymmetry in neuromelanin sensitive contrast might also have limited the sensitivity to detect a between group difference in the left LC.

Patients were taking their usual medication during clinical and neuropsychological testing. While this reduced the confounding effects of motor impairment on the test outcomes, dopaminergic treatment may have influenced orthostatic regulation. Hence, orthostatic blood pressure measurements might have been different off dopaminergic treatment. Similarly, antihypertensive treatment was not withdrawn prior to orthostatic blood pressure measurements. Excluding patients on antihypertensive treatment did not influence our conclusions regarding the association between LC contrast and orthostatic systolic blood pressure change.

### Conclusion

Our results show that neuromelanin sensitive MTw MRI at ultra-high field strength can map spatial gradients of structural disintegration in the LC, showing a preferential involvement of the caudal portion of LC in PD and a clear relationship between reduced structural integrity of caudal LC and orthostatic dysregulation and apathy.

## Data Availability

The data supporting the findings of this study is available from the corresponding author upon reasonable request.

## (6) Acknowledgments

We thank Dr. Rong Ye and Professor James B. Rowe from the Department of Clinical Neurosciences, University of Cambridge and Cambridge University Hospitals NHS Foundation Trust for kindly sharing their probabilistic atlas of the locus coeruleus and for providing support in setting up the image post-processing pipeline. We would also like to thank the patients and healthy participants who participated in our study.

The study was funded by the Independent Research Fund Denmark (Grant No. 7016-00226B), The Novo Nordisk Foundation (Grant No. NNF16OC0023090) and the Danish Parkinson Association (Grant No. A71 & A289). The 7T scanner was donated by the John and Birthe Meyer Foundation and The Danish Agency for Science, Technology and Innovation (grant no. 0601-01370B).

HS holds a 5-year clinical professorship with focus on in precision medicine at the Faculty of Health Sciences and Medicine, University of Copenhagen which is sponsored by the Lundbeck Foundation (Grant No. R186-2015-2138). AL was supported by the Danish Parkinson Associtation. HS and SF were supported by the Novo Nordisk Foundation Synergy Grant (Grant No. NNF17OC0027872,UHeal).

## (7) Authors’ Roles

1. Research project: A. Conception, B. Organization, C. Execution;
2. Statistical Analysis: A. Design, B. Execution, C. Review and Critique;
3. Manuscript: A. Writing of the first draft, B. Review and Critique.

CM: 1A, 1B, 1C, 2A, 2B, 3A

DM: 1A, 1B, 1C, 2A, 2C, 3B

SF: 2A, 2B, 2C, 3B

MM: 2B, 3B

VB: 1C, 2C, 3B

KM: 2A, 2B, 2C, 3B

EP: 1C, 2C, 3B

AH: 2C, 3B

AL: 1A, 2C, 3B

HS: 1A, 1B, 2C, 3B

## (8) Financial Disclosures of all authors (for the preceding 36 months)

HS has received honoraria as speaker from Sanofi Genzyme, Denmark and Novartis, Denmark, as consultant from Sanofi Genzyme, Denmark, Lophora, Denmark, Lundbeck AS, Denmark, and as editor-in-chief (Neuroimage Clinical) and senior editor (NeuroImage) from Elsevier Publishers, Amsterdam, The Netherlands. He has received royalties as book editor from Springer Publishers, Stuttgart, Germany and from Gyldendal Publishers, Copenhagen, Denmark.

AL has received honoraria as speaker from AbbVie, United States and GE Healthcare, Denmark. The other authors declare no conflict of interest.

## (12) Supplementary figures

**Supplementary figure 1.**
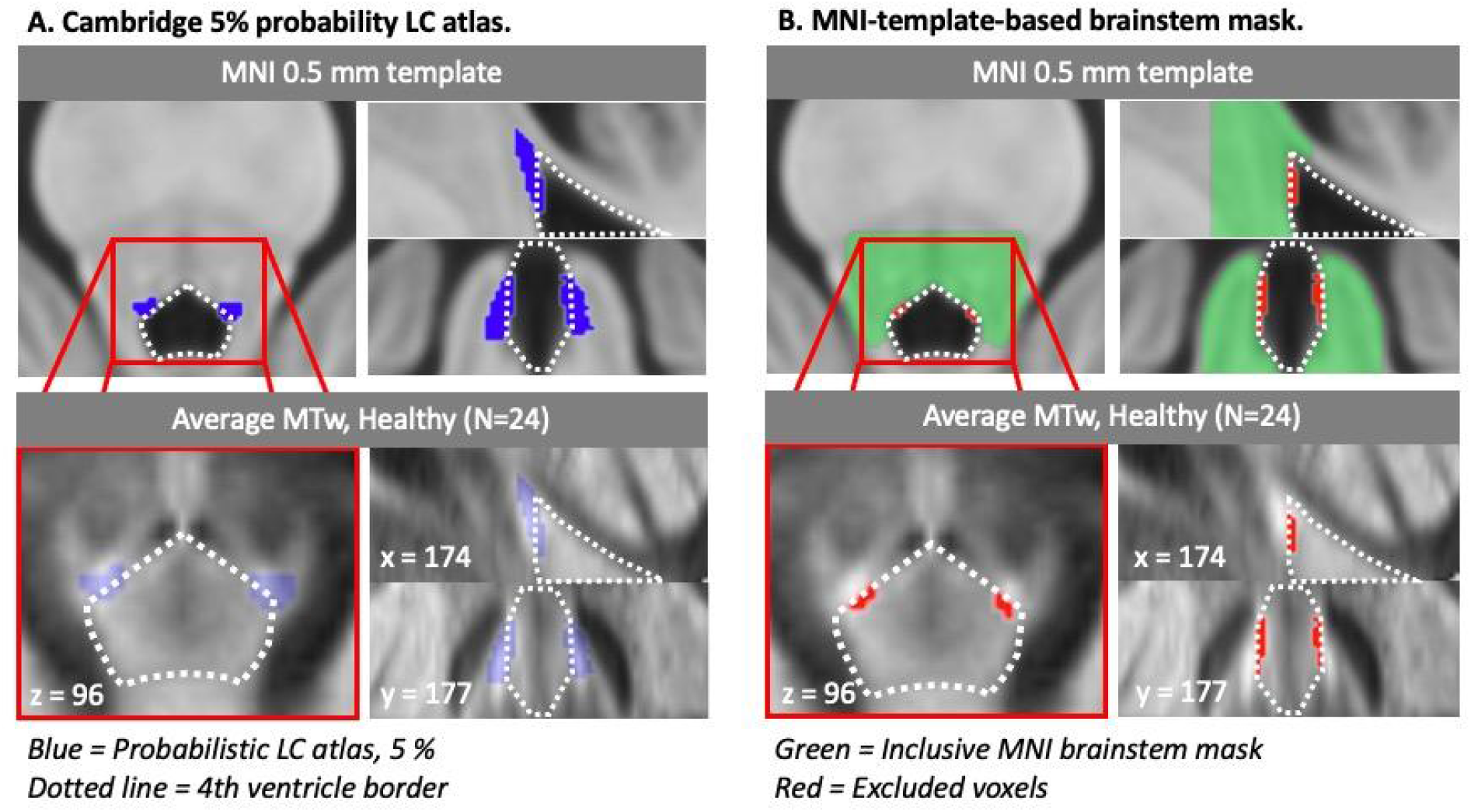
LC region of interest. An issue for extracting LC ROI statistics was the hyperintense border of the 4th ventricle on MT-weighted images (bottom panels, dashed line). By averaging MT-weighted images from all HCs, we identified a non-negligeable overlap between the Cambridge 5% probability LC atlas (blue) and hyperintense voxels belonging to the 4th ventricle border. We therefore restricted the LC ROI by creating a threshold-based brainstem mask from the MNI template (green), which excluded voxels in the immediate vicinity of the 4th ventricle (red) (B).

**Supplementary figure 2.**
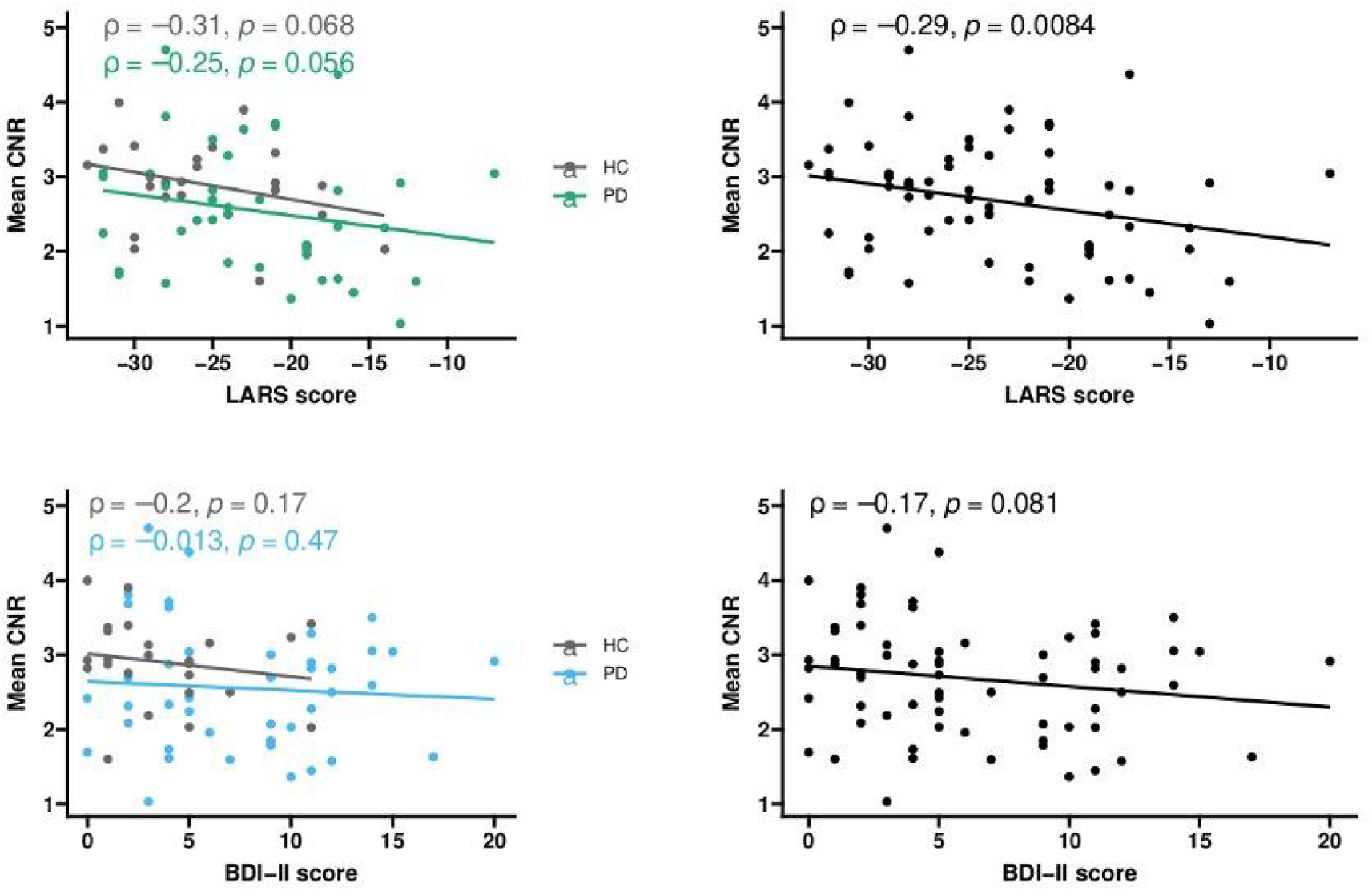
Correlations between overall LC CNR_MTw_ and apathy and depression scores in all participants. Apathy and depression was assessed in all participants. We evaluated whether adding overall LC CNR_MTw_ values of healthy participants would increase sensitivity in detecting correlations between LC CNR_MTw_ and the severity of these features. Pooling the data from both groups revealed a highly significant correlation between apathy scores and overall LC CNR_MTw_, while there was no significant correlation for depression scores.

**Supplementary figure 3.**
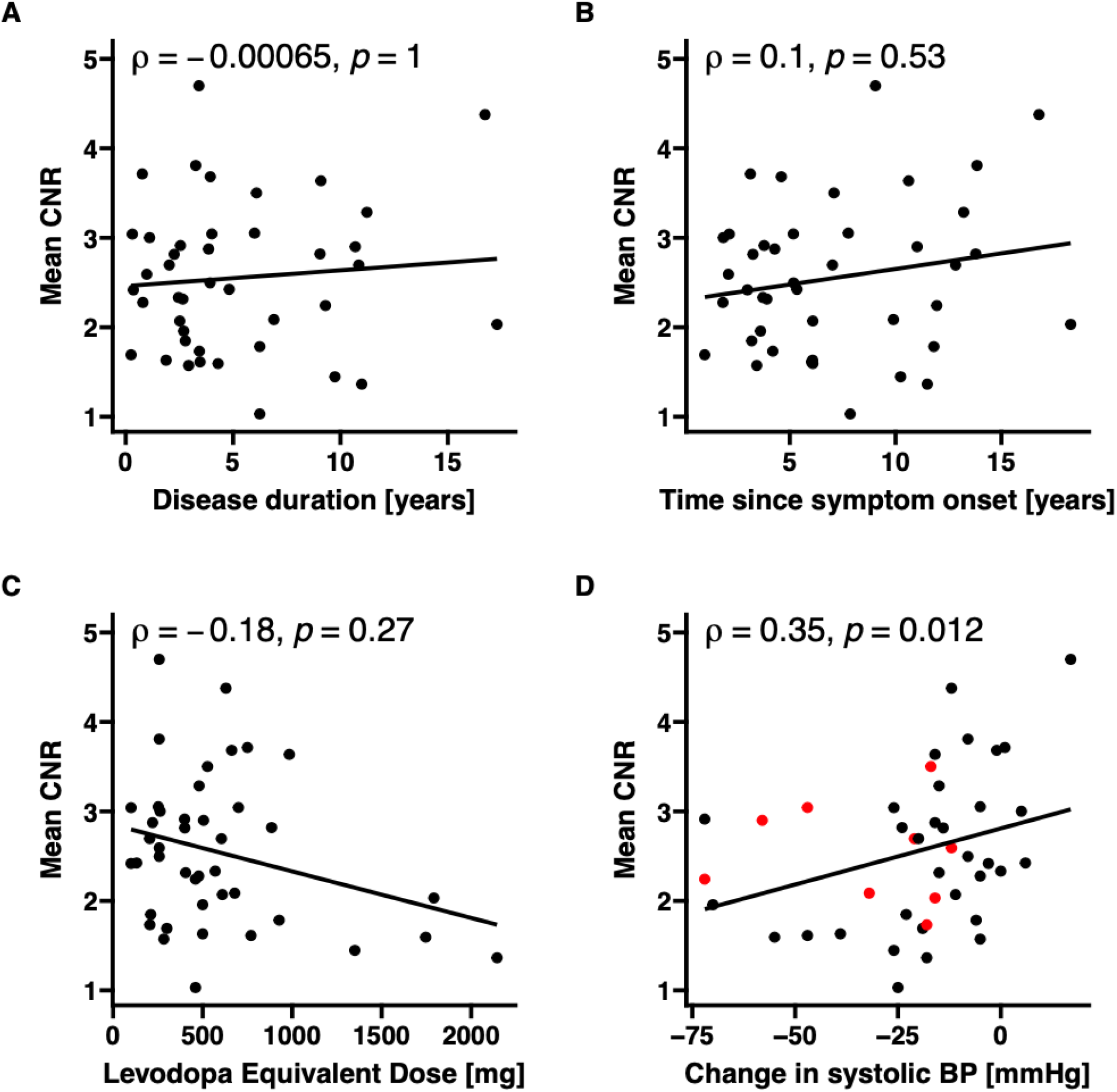
Correlations with disease duration, time since symptom onset, and Levodopa Equivalent Dose and effect of antihypertensive treatment. No correlations were found between LC CNR_MTw_ and disease duration (A), time since symptom onset (B) or Levodopa Equivalent Dose (C). The inclusion of patients on antihypertensive treatment did not influence the correlation between LC CNR_MTw_ and orthostatic change in systolic blood pressure (D, red dots = patients on antihypertensive treatment).

**Supplementary figure 4.**
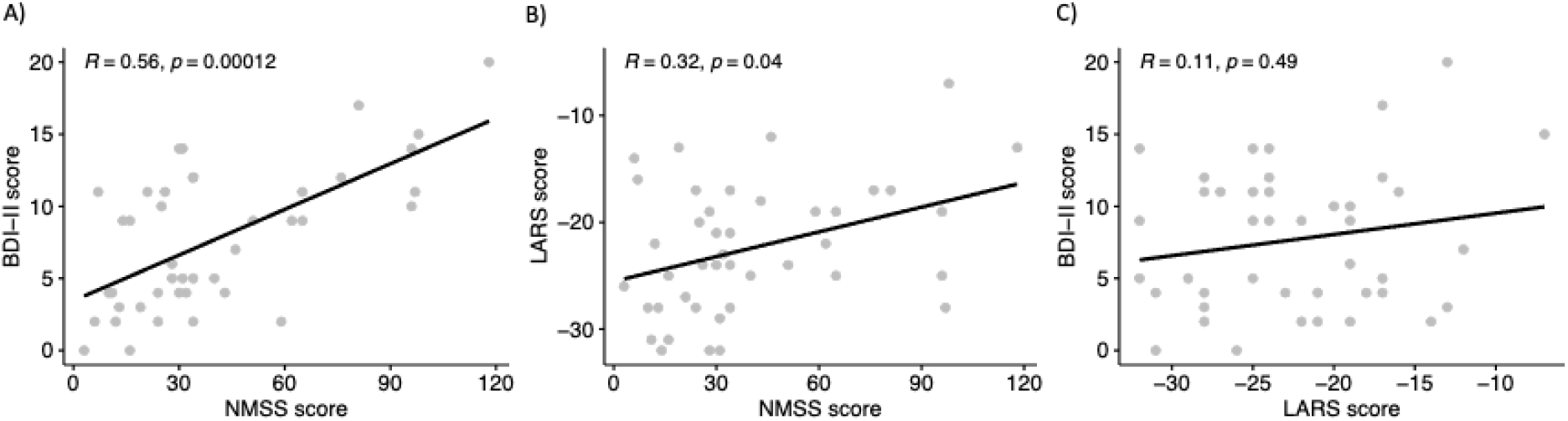
Correlations of non-motor symptoms. Non-motor symptom scale (NMSS) scores correlated positively with Beck’s Depression Inventory II (BDI-II) scores (A), and Lille Apathy rating scale (LARS) scores (B). LARS & BDI-II scores were not significantly correlated. Correlations were assessed using Spearman Rank correlation.

## Notes

### Competing Interest Statement

All authors have completed the ICMJE uniform disclosure form at www.icmje.org/coi_disclosure.pdf and declare: HS, CM, DM and MM had financial support from the Independent Research Fund Denmark (Grant No. 7016-00226B), The Novo Nordisk Foundation (Grant No. NNF16OC0023090) and the Danish Parkinson Association (Grant No. A71 & A289) for the submitted work; HS has received research grants from the Lundbeck Foundation (Grant No. R186-2015-2138) and the Novo Nordisk Foundation Synergy Grant (Grant No. NNF17OC0027872,UHeal) and has received honoraria as speaker from Sanofi Genzyme, Denmark and Novartis, Denmark, as consultant from Sanofi Genzyme, Denmark, Lophora, Denmark, Lundbeck AS, Denmark, and as editor-in-chief (Neuroimage Clinical) and senior editor (NeuroImage) from Elsevier Publishers, Amsterdam, The Netherlands; HS has received royalties as book editor from Springer Publishers, Stuttgart, Germany and from Gyldendal Publishers, Copenhagen, Denmark; AL has received honoraria from AbbVie, United States and GE Healthcare, Denmark, and support from the Danish Parkinson Association; SF was supported by the Novo Nordisk Foundation Synergy Grant (Grant No. NNF17OC0027872,UHeal); EP is treasurer for the Danish Society for Medical Magnetic Resonance; no other relationships or activities that could appear to have influenced the submitted work.

### Clinical Trial

NCT03866044

### Author Declarations

The Regional Committee on Health Research Ethics of the Capital Region of Denmark (Record-id: H-18021857)

